# The association of household food insecurity and HIV infection with common mental disorders among newly diagnosed tuberculosis patients in Botswana

**DOI:** 10.1101/2020.08.15.20175315

**Authors:** Qiao Wang, Mbatshi Dima, Ari Ho-Foster, Keneilwe Molebatsi, Chawangwa Modongo, Nicola M. Zetola, Sanghyuk S. Shin

## Abstract

**Objective:** To determine the association between food insecurity and HIV-infection with depression and anxiety among new tuberculosis patients.

**Design:** Our cross-sectional study assessed depression, anxiety, and food insecurity with Patient Health Questionnaire (PHQ9), Zung Anxiety Self-Assessment Scale (ZUNG), and Household Food Insecurity Access Scale, respectively. Poisson regression models with robust variance were used to examine correlates of depression (PHQ9 ≥ 10) and anxiety (ZUNG ≥ 36)

**Setting:** Gaborone, Botswana.

**Participants:** Patients who were newly diagnosed with tuberculosis.

**Results:** Between January and December 2019, we enrolled 180 TB patients from primary health clinics in Botswana. Overall, 99 (55.0%) were HIV-positive, 47 (26.1%),85 (47.2%), and 69 (38.5%) indicated depression, anxiety, and moderate to severe food insecurity, respectively. After adjusting for potential confounders, food insecurity was associated with a higher prevalence of depression (adjusted prevalence ratio [aPR] =2.30; 95% confidence interval [CI] = 1.40, 3.78) and anxiety (aPR = 1.41; 95% CI = 1.05, 1.91). Prevalence of depression and anxiety were similar between HIV-infected and -uninfected participants. Estimates remained comparable when restricted to HIV-infected participants.

**Conclusions:** Mental disorders may be affected by food insecurity among new tuberculosis patients, regardless of HIV status.

## Introduction

Mental illness is a leading cause of disability, with approximately 970 million people being affected worldwide. (1) Compared with the general population, individuals affected by tuberculosis (TB) are significantly more likely to have poor mental health due to biological and social complexities. (2) Common mental illness, including depression and anxiety disorders, imposes immense costs on healthcare systems and poor quality of life on patients, and can be important barriers for TB diagnosis and treatment. Moreover, depression may be associated with unfavorable TB treatment outcomes due to medication nonadherence, (3) which has been cited as the fundamental obstacle in effective TB control. (4) Therefore, mental health is an integral part of achieving WHO’s The End TB Strategy and eliminating the global tuberculosis epidemic. (5) Unfortunately, the majority of TB and mental illness occur in low- and middle-income countries (LMIC), (6) where mental health is often overlooked due to myriad challenges such as lack of mental health specialists, limited treatment options, and other competing public health priorities. (6)

Meanwhile, individuals with TB often experiences malnutrition (7), which may be exacerbated by food insecurity. Globally, about 2 billion people suffer from moderate to severe levels of food insecurity. (8) According to The United States Department of Agriculture, a household is defined as food insecure if at least one member experiences insufficient or inconsistent access to nutritionally adequate and safe food. (9) A survey of patients with TB/HIV co-infection in multiple African cities reported that only 10% TB patients feel they are food secure. (10) Food insecurity not only negatively affects general physical health, but also has important ramifications for adverse mental health. For instance, studies have shown that adults with food insecurity are more likely to have anxiety disorder, depression, and suicidal ideation compare to those who are food secure. (11)

Globally, Botswana has one of the highest TB incidence – 300 cases per 100,000 population in 2017. (12) About 60% of all TB patients also have comorbid HIV infection, (12) which has been suggested as a risk factor in psychiatric populations. (13, 14) People living with HIV/AIDS may have an increased risk of developing mood disorders and clinical depressive symptoms triggered by distress associated with initial HIV diagnosis, disease progression, difficult life circumstances, hospitalization, HIV-induced brain injury, and adverse effects of antiretroviral medications. (14) The most recent data indicate that psychiatric morbidity in the country contributes an estimated 4.6% of the global burden of disease. (15) There are 0.5 mental health outpatient facilities, 2.5 psychiatrists, and 15.2 psychologists available per million population in spite of the heavy burden of psychological distress. (15)

For the past few decades, Botswana has been experiencing rapid urbanization and positive economic growth with a significant decline in poverty incidence. (16) At the same time, many challenges remain, including food insecurity. The Food and Agricultural Organization (FAO) indicated that the average prevalence of severe food insecurity was 41.3% between 2016 and 2018. (17) It is crucial to understand the relationship between food insecurity and mental health among the socially vulnerable TB patients because, while the expansion and integration of mental health care can be a challenging and gradual task in resource-limited settings, social support programs, particularly food supplementation, may be relatively low-intensity and straightforward for policy delivery and implementation. The objectives of our study were to examine the prevalence of depression and anxiety among newly diagnosed TB patients in Botswana, and to explore the association between those conditions with food insecurity and HIV co-infection.

## Methods

### Study design, population, and eligibility

We conducted a cross-sectional study and recruited participants with newly diagnosed pulmonary or extrapulmonary TB from primary healthcare clinics in Gaborone, Botswana. Participants were deemed eligible if they were at least 18 years old with a verified TB diagnosis, and were recruited by our research staff at the time of receiving TB diagnosis. According to national guidelines during the study period, the first-line diagnostic test for TB was sputum smear microscopy or GeneXpert MTB/RIF for patients who experienced prolonged general pulmonary TB symptoms such as cough,fever, shortness of breath, night sweats, weight loss, and hemoptysis. (18) Patients with indications for culture testing, such as symptomatic individuals at high risk of multidrug106 resistant TB (MDR-TB), also underwent examination by mycobacterial culture and drug susceptibility testing (DST) with the Mycobacteria Growth Indicator Tube (MGIT) method. A chest radiography was performed in symptomatic individuals who had negative sputum smears or negative Xpert MTB/RIF. For patients who had negative results in all testing, TB diagnosis was made by physician based on clinical presentations and/or lymph node examinations. A rapid HIV test was conducted among participants who have not had an HIV test done before, or for those who have had a negative result in previous testing.

### Data collection

Socio-demographic data were collected via face-to-face interview and included age, gender, education, monthly income, marital status, smoking, and alcohol history. We also collected clinical information that included TB symptoms, duration of the symptoms, HIV test result and HIV diagnosis date if any, anti-retroviral treatment (ART) history, and results of smear microscopy, Xpert MTB/RIF, mycobacterial culture, and chest X-ray. CD4 T-cell data closest to the time of enrollment were extracted and integrated from the national HIV database for HIV-infected participants.

We administered standardized questionnaires to assess the presence and severity of depressive symptoms, anxiety symptoms, and household food insecurity of enrolled participants. For simplicity, we will use depression and anxiety to describe depressive symptoms and anxiety symptoms, respectively, hereinafter.

Depression was assessed by the Patient Health Questionnaire (PHQ-9), a 9-item self-report tool that screened depression as participants identified how much each item statement applied to them within the past two weeks. (19) Examples of item statements include “feeling down, depressed or hopeless” and “little interest or pleasure in doing things”. Answer choices were scored on a Likert-type scale that ranged from “not at all”,several days”, “more than half the days”, to “nearly every day”. PHQ-9 has been validated in multiple sub-Saharan African countries to assess depression, including Botswana. (20)

Anxiety was assessed by Zung Self-Rating Anxiety Scale (ZUNG), a 20-item self-report scale that measured anxiety levels.(21) Similar to PHQ-9, participants indicated how much each of the 20 statements applied to them within the past several days, and replies included “none or a little of the time”, “some of the time”, “good part of the time”, and “most or all of the time”. Examples of item statements include “I feel afraid for no reason at all” and “I get upset easily or feel panicky”.

Finally, food insecurity was evaluated by the Household Food Insecurity Access Scale (HFIAS), a 9-item questionnaire widely used across different culture, including Botswana, (16) to distinguish food insecure households and to determine the degree of food insecurity. (22) Examples on this questionnaire include “In the past four weeks, did you worry that your home would not have enough food?” and “In the past four weeks, did you or anyone in the house go to sleep at night hungry because there was not enough food?” Answer choices were scored on a Likert-type scale that ranged from “never”, “rarely (1 or 2 times)”, “sometimes (3-10 times)”, to “often (more than 10 times)”.

All questionnaires were prepared in English and Setswana, both official languages of Botswana. Participants were able to pick the language of preference during interview. All participants with PHQ-9 scores indicative of depression and suicidal ideation were provided with appropriate psychiatric referrals.

### Statistical analysis

The outcomes of interest for this study were the prevalence of depression and anxiety among newly diagnosed TB patients, and the roles of food insecurity and HIV status on a) depression, and b) anxiety. Patients were categorized as having depression if they scored 10 points or more on PHQ-9, (19) and having anxiety if they scored 36 points (raw score) or more on ZUNG. (21) Prevalence estimates of depression and anxiety were calculated as the number of observed participants with depression and anxiety, respectively, divided by the total number of enrolled participants, and the 95% confidence limits were obtained by the Wilson score method. Based on HFIAS scores, participants were categorized as “food secure”, “mildly insecure”, “moderately insecure”, or “severely insecure” households. (22) For the purpose of analysis, we dichotomized participants as experiencing “food security or mild insecurity”, or “moderate to severe insecurity”. (23) Monthly income was categorized as having “no income” or “some income”. As recommended for estimating prevalence ratios in cross-sectional studies where the outcome of interest was common, Poisson regression models with robust variance were used to examine the bivariate associations between selected covariates and each of the mental disorder outcomes. (24) Independent covariates were selected based on a priori knowledge of predictors of adverse mental health, which include age, gender, HIV status, monthly income, and food insecurity. All covariates were included in the multivariable model regardless of the observed bivariate associations. Subsequently, we analyzed the bivariate associations between age, gender, monthly income, ART status, CD4 T-cell count, food insecurity and each outcome among participants infected with HIV. Similarly, multivariable model was constructed to determine the independent effect of each covariate, regardless of their bivariate associations. However, because this is a smaller subset of participants, we present the model including only food insecurity, ART status, and CD4 T-cell count in this paper. Estimates were similar between the smaller model and the full model.

Data analysis was carried out in SAS version 9.4 (SAS Institute, Cary, North Carolina, U.S.). In accordance with recent guidelines, no alpha cutoff was specified for statistical significance. (25)

### Ethical considerations

This study was approved by the institutional review board (IRB) at University of California, Irvine, and the Human Research Development Committee of the Botswana Ministry of Health. Written informed consent was obtained from all participants prior to enrollment.

## Results

Between January and December of 2019, 180 participants were recruited into the study. Overall, 99 (55.0%) were HIV positive, 64 (35.6%) were female, 155 (86.1%) were single, 137 (76.1%) were diagnosed with pulmonary TB, 17 (9.4%) had no formal education, and 65 (36.1%) had no income (Table 1). Among those who were HIV co-infected, the median duration since HIV diagnosis was 43 months, median CD4 T-cell count was 335, and 31 (31.3%) had never taken ART, of whom 90% were newly diagnosed with HIV at or within one month of study enrollment. Over half of all participants reported being food secure, and 15 (8.4%), 17 (9.5%), 52 (29.0%) reported experiencing mild insecurity,moderate insecurity, and severe insecurity, respectively (Table 1).

**Table 1.**
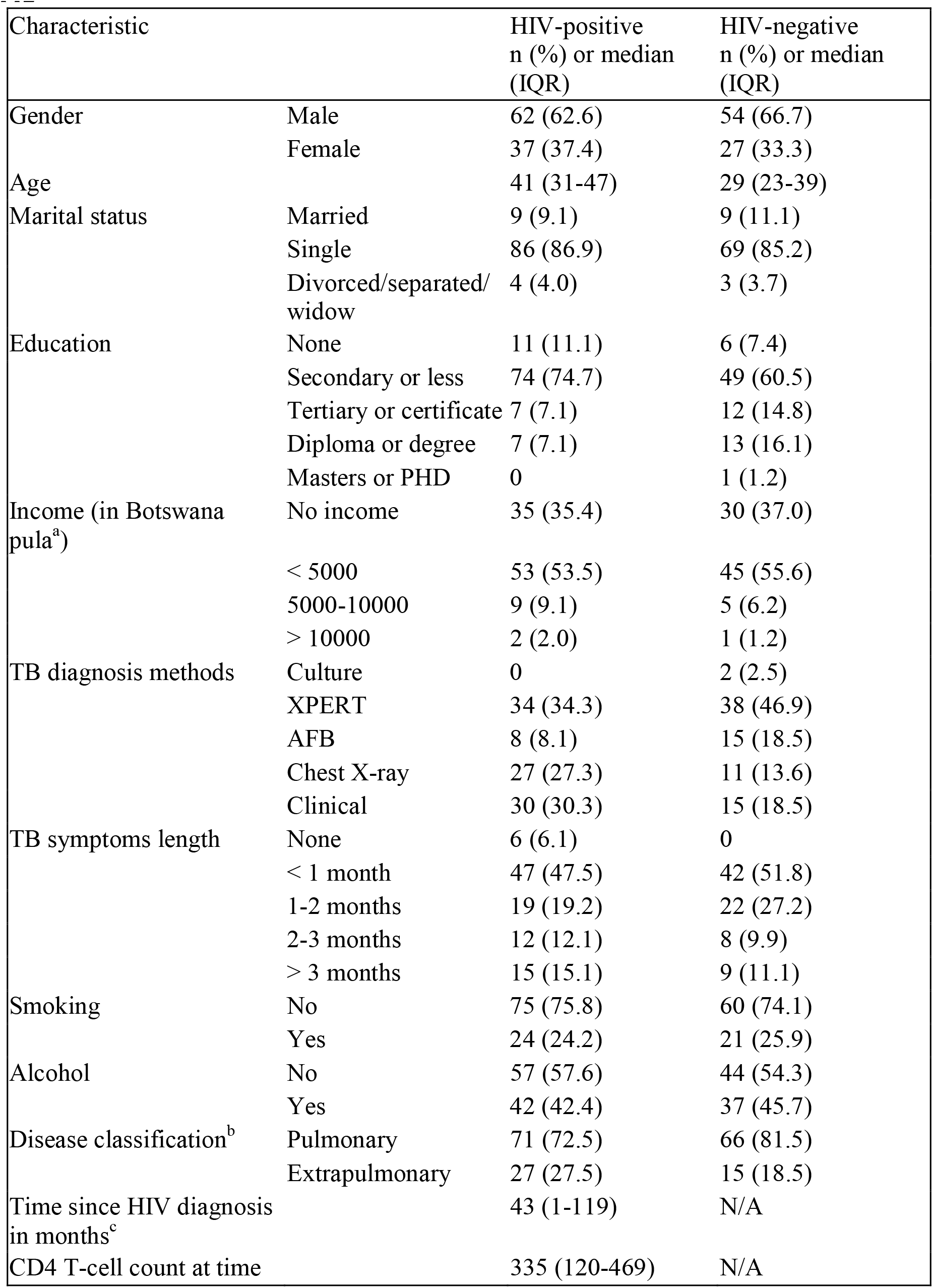
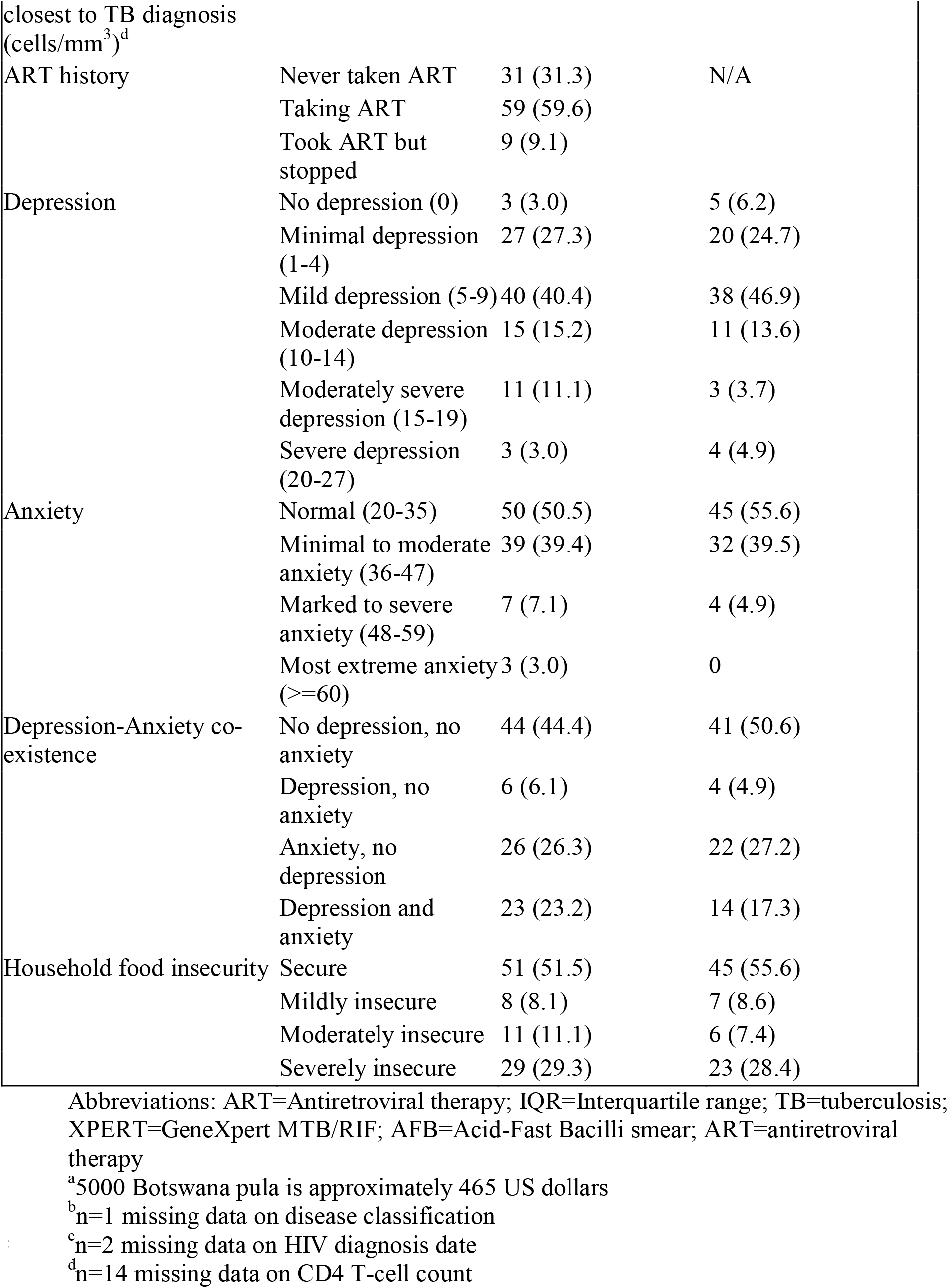
Characteristics of study participants in Botswana, 2019 (n =180)

The overall prevalence of depression, anxiety, and co-existence of both conditions were 26.1%, 47.2%, and 20.6%, respectively. Among those who were depressed, 26 (14.4%), 14 (7.8%), and 7 (3.9%)had moderate, moderately severe, and severe depression, respectively (Table 1).Twenty-seven (15.1%) participants had PHQ-9 scores indicative of suicidal ideation. Additionally, 71 (39.4%), 11 (6.1%), and 3 (1.7%) had minimal to moderate, marked to severe, and most extreme anxiety, respectively.

Table 2 shows the results of Poisson regression analysis of covariates associated with depression. In bivariate analysis, moderate to severe food insecurity was associated with a higher prevalence of depression (crude prevalence ratio [cPR] = 2.37; 95% confidence interval [CI] = 1.44, 3.91). After adjusting for age, gender, monthly income, and HIV co-infection status, the association between moderate to severe food insecurity and depression remained similar (adjusted prevalence ratio [aPR] = 2.30; 95% CI = 1.40,3.78).

**Table 2.** Correlates of depression^a^ among study participants in Botswana, 2019 (n = 180)

| Variables |  | Prevalence<br>n/N (%) | Crude PR<br>(95% CI) | Adjusted PR<br>(95% CI) |
| --- | --- | --- | --- | --- |
| Age |  | N/A | 1.01 (0.99, 1.03) | 1.01 (0.99, 1.02) |
| Gender | Male | 26/116 (22.4) | 1.00 | 1.00 |
|  | Female | 21/64 (32.8) | 1.46 (0.90, 2.38) | 1.41 (0.88, 2.27) |
| HIV | Negative | 18/81 (22.2) | 1.00 | 1.00 |
|  | Positive | 29/99 (29.3) | 1.32 (0.79, 2.19) | 1.20 (0.73, 1.98) |
| Income | No | 19/65 (29.2) | 1.00 | 1.00 |
|  | Yes | 28/115 (24.4) | 0.83 (0.51, 1.37) | 0.95 (0.59, 1.52) |
| Food insecurity | None or mildly insecure | 19/111 (17.1) | 1.00 | 1.00 |
|  | Moderate to severe insecure | 28/69 (40.6) | **2.37 (1.44, 3.91)** | **2.30 (1.40, 3.78)** |

Abbreviations: PR=Prevalence ratio; CI=Confidence interval
<sup>a</sup>The presence of depression is defined as scoring equal to or more than 10 points on the Patient Health Questionnaire (PHQ-9).

Similar findings were observed for the outcome of anxiety; moderate to severe food insecurity was associated with a higher prevalence of anxiety (cPR = 1.43; 95% CI = 1.06, 1.93; Table 3). The association remained robust in multivariable Poisson analysis after adjusting for age, gender, monthly income, and HIV co-infection status (aPR = 1.41; 95% CI = 1.05, 1.91; Table 3).

**Table 3.** Correlates of anxiety^a^ among study participants in Botswana, 2019 (n = 180)

| Variables |  | Prevalence<br>n/N (%) | Crude PR<br>(95% CI) | Adjusted PR<br>(95% CI) |
| --- | --- | --- | --- | --- |
| Age |  | N/A | 1.01 (0.99, 1.02) | 1.01 (0.99, 1.02) |
| Gender | Male | 52/116 (44.8) | 1.00 | 1.00 |
|  | Female | 33/64 (51.6) | 1.15 (0.84, 1.57) | 1.14 (0.84, 1.56) |
| HIV | Negative | 36/81 (44.4) | 1.00 | 1.00 |
|  | Positive | 49/99 (49.5) | 1.11 (0.81, 1.53) | 1.05 (0.77, 1.42) |
| Income | No | 32/65 (49.2) | 1.00 | 1.00 |
|  | Yes | 53/115 (46.1) | 0.94 (0.69, 1.30) | 0.98 (0.72, 1.34) |
| Food insecurity | None or mildly<br>insecure | 45/111 (40.5) | 1.00 | 1.00 |
|  | Moderate to severe<br>insecure | 40/69 (58.0) | **1.43 (1.06, 1.93)** | **1.41 (1.05, 1.91)** |

Abbreviations: PR=Prevalence ratio; CI=Confidence interval
<sup>a</sup>The presence of anxiety is defined as scoring equal to or more than 36 points (raw score) on the Zung Anxiety Self-Assessment Scale (ZUNG).

Among HIV-co-infected participants, the estimates were similar in the bivariate analysis for depression (cPR = 2.41; 95% CI = 1.28, 4.55; Table 4) and anxiety (cPR = 1.54; 95% CI = 1.04, 2.27). Accounting for ART status and CD4 count closest to the time of TB diagnosis, food insecurity continued to be associated with increased symptoms of depression (aPR = 2.33; 95% CI = 1.24, 4.38) and anxiety (aPR = 1.53; 95% CI = 1.03, 2.26) in the multivariable model.

**Table 4.** Correlates of depression^a^ and anxiety^b^ among HIV-infected participants in Botswana, 2019 (n = 99)

|  |  | Depression |  | Anxiety |  |
| --- | --- | --- | --- | --- | --- |
| Variables |  | Crude PR<br>(95% CI) | Adjusted PR<br>(95% CI) | Crude PR<br>(95% CI) | Adjusted PR<br>(95% CI) |
| Age |  | 1.01 (0.99, 1.04) | Not included | 1.01 (0.99, 1.03) | Not included |
| Gender | Male | 1.00 | Not included | 1.00 | Not included |
|  | Female | 1.56 (0.86, 2.86) |  | 1.16 (0.78, 1.72) |  |
| Income | No | 1.00 | Not included | 1.00 | Not included |
|  | Yes | 0.67 (0.37, 1.23) |  | 0.86 (0.58, 1.29) |  |
| ART | No | 1.00 | 1.00 | 1.00 | 1.00 |
|  | Yes | 1.29 (0.67, 2.47) | 1.14 (0.58, 2.21) | 1.17 (0.77, 1.78) | 1.12 (0.73, 1.73) |
| CD4 T-cell<br>count<br>(cells/mm<sup>3</sup>) | ≤ 200 | 1.00 | 1.00 | 1.00 | 1.00 |
|  | > 200 | 1.48 (0.70, 3.11) | 1.26 (0.59, 2.72) | 1.17 (0.72, 1.91) | 1.06 (0.64, 1.75) |
|  | Missing | 0.92 (0.28, 3.03) | 0.87 (0.26, 2.94) | 1.32 (0.72, 2.43) | 1.27 (0.68, 2.35) |
| Food<br>insecurity | None or mildly<br>insecure | 1.00 | 1.00 | 1.00 | 1.00 |
|  | Moderate to<br>severe insecure | **2.41 (1.28, 4.55)** | **2.33 (1.24, 4.38)** | **1.54 (1.04, 2.27)** | **1.53 (1.03, 2.26)** |

Abbreviations: PR=Prevalence ratio; CI=Confidence interval; ART=antiretroviral therapy
<sup>a</sup>The presence of depression is defined as scoring equal to or more than 10 points on the Patient Health Questionnaire (PHQ-9).
<sup>b</sup>The presence of anxiety is defined as scoring equal to or more than 36 points (raw score) on the Zung Anxiety Self-Assessment Scale (ZUNG).

## Discussion

In our population of TB patients, we showed that household food insecurity was the most important factor associated with symptoms of depression and anxiety, independent of HIV co-infection status, age, monthly income, and gender. Our findings confirmed previous research that showed a linkage between food insecurity and poor mental health, (26, 27) and extended this finding in TB patients. Several reasons may explain this association. First, as a consequence of food insecurity, deficiency in critical micronutrient could lead to depression and anxiety via biological pathways. (28) Additionally, food insecurity encompasses not only hunger and under-nutrition, but also uncertainty and distress over the access of food. Thus, food insecurity may compromise mental health by generating instability and unpredictability into lives of the already socioeconomically vulnerable TB population. (26) This pathway could be independent of the degree of social and economic deprivation, as a global analysis of 149 countries concluded an association between food insecurity and mental illness in a dose-response fashion, after controlling for socioeconomic status. (27)

To our knowledge, this is the first study that examined the association between food insecurity and mental illness among new TB patients in a high burden setting. In compliance with The End TB Strategy, Botswana has social (e.g. food support) programs to help TB-affected individuals improve nutritional status and treatment adherence, and alleviate the steep costs associated with the disease and treatment. Specifically, TB patients with confirmed diagnosis may receive food allocations that include cooking oil, beans, and maize flour with additive vitamins. However, the food rations are usually not enough to support every member of the household. In addition, TB renders many patients unable to work and contribute to household expenditures, and caregivers may still experience household food insecurity as they face challenges of providing for their family due to income loss. (29) The total financial costs of TB can be catastrophic for patients – up to 58% of annual individual income and 39% of household income in LMIC, and income loss constitute the largest portion of all TB-related cost at 60%. (29) The cyclical nature of the relationship between food insecurity and TB fosters sustained impoverishment and makes it difficult for household to break the cycle without outside intervention. Malnutrition, a consequence of food insecurity, may predispose patients for risks of TB infection and TB disease reactivation. (30)

Comparable to national data released by FAO, (17) our survey indicated 38.5% of the participants had experienced moderate to severe food insecurity prior to receiving their TB diagnosis. Further, a separate survey estimated that only 12% of households report to be “food secure” in urban parts of Gaborone. (16) These findings highlight the pervasive food insecure households among TB patients and those at risk for TB in Botswana, which lend support for food assistance programs and social protection interventions to target populations who are at high risks for TB. Other research suggests members of households who participate in food supplementation programs have lower rates of psychological distress; by reducing food insecurity, these programs can be effective in improving mental health and overall security among vulnerable populations. (31)

In this study, we revealed that anxiety and depression were highly prevalent among patients newly diagnosed with TB in Botswana. Approximately 26% of enrolled TB patients had PHQ-9 scores indicative of moderate to severe depression and 15% indicative of suicidal ideation. Our estimate was comparable to recent studies in several countries that examined depression (moderate to severe) among TB patients, including Ethiopia (17.6 - 54.0%), (32-34) South Africa (32.9%), (13) Angola (49.4%), (35) and China (18.1%). (36) They were, however, considerably lower than those found in Ethiopia using PHQ-9 by Ambaw *et al*.; (32) these differences may have arisen from variations in study location and population. For example, roughly 45% of the participants in the Ethiopian study had no formal education and over half were married.

Approximately 46% of our participants reported anxiety symptoms, comparable to published estimates of other settings: Ethiopia (41.5%), (34) Angola (38.3%), (35) and China (18.4%). (36) None of those studies utilized the ZUNG scale; instead, Generalized Anxiety Disorder Questionnaire (GAD-7) and Hospital Anxiety and Depression Scale (HADS) were used to assess the presence of anxiety symptoms, which might be an explanation for the discrepant results. Our estimate was much higher than those published by Wang *et al*. of Chinese TB patients. (36) One explanation for this discrepancy could be patient characteristics: in the Chinese study over half of participants had tertiary or higher education levels; majority were married; and patients with extrapulmonary TB were excluded.

We did not find an association between HIV comorbidity and mental disorders in our sample. This finding is in agreement with some studies, (32, 33) but not others. (13,34) Studies that reported an association between comorbidity and mental disorders have posited HIV-associated stigma as the plausible driving force. (13, 34) Among people with TB, factors such as malnutrition, poor physical health, socioeconomic adversities, and interaction between these factors may have a stronger influence on depression and anxiety than HIV-related stigma. Furthermore, following WHO’s guidelines in 2015, nearly all sub-Saharan African countries adopted universal HIV care policies irrespective of CD4 T-cell count (‘treat all’), including Botswana. The well-developed, universal access to HIV care may have also influenced mental health in our population of TB patients. Individuals who were co-infected but not on ART predominantly discovered their HIV status at enrollment or within one month of study enrollment, and further research is needed among this group as HIV-related stigma may be perceived differently in these individuals.

We note several limitations associated with our research. First, depression and anxiety assessment relied on self-reporting using screening questionnaires. This may lead to self-report bias, particularly under-reporting owing to stigmatization of psychological disorders. However, we tried to minimize this by training the research staff on administering the questionnaires with consistency and without judgment, and by asking the participants to record their answer on the tablet directly. Second, the PHQ-9 and ZUNG screening tools have not been validated in Botswana, nor in TB patients in Botswana. The English version of these questionnaires were translated by a native speaker of Setswana. Some items on the questionnaires overlap with TB symptoms, including fatigue, loss of appetite, and insomnia, which may lead to misclassification of depressive and anxiety cases. As a consequence, our estimate of a\ssociations between depression, anxiety, and independent variables may be biased toward the null. Finally, due to the cross-sectional study design, statistical associations may not be representative of casual relationships between exposure and outcome variables.

In conclusion, we have shown that common mental disorders are highly prevalent and food insecurity is associated with poor mental health among new TB patients. This study addresses the critical need for integration of TB, mental health, and social services (i.e. food assistance) in Botswana. Current programs intended to support individuals struggling with food security may require additional measures to ensure access to and adequacy of nutritious and sufficient food for the household. Given the global goal of ending TB and rising interest in mental health, longitudinal research is needed to determine the temporal relationship between food insecurity and mental disorders, and to identify potential pathways through which food insecurity may elevate the risk of common mental disorders among TB patients.

## Data Availability

The data that support the findings of this study are available on request from the corresponding author, S.S. Shin. The data are not publicly available due to their containing information that could compromise the privacy of research participants.

## Acknowledgements

We are thankful to the study participants who made this study possible. The authors also acknowledge the following sources of support:

## Financial Support

This work was supported by the National Institute of Allergy and Infectious Diseases [grant number K01AI118559]. The funding source had no role in the design, analysis or writing of this article.

## Conflicts of Interest

None.

## Notes

### Competing Interest Statement

The authors have declared no competing interest.

### Author Declarations

University of California, Irvine IRB and Human Research Development Committee of the Botswana Ministry of Health

